# Mechanical hyperalgesia and neuropathic pain qualities impart risk for chronic postoperative pain after total knee replacement

**DOI:** 10.1101/2024.01.16.24301372

**Authors:** Andrew D. Vigotsky, Olivia Cong, Camila B Pinto, Joana Barroso, Jennifer Perez, Kristian Kjaer Petersen, Lars Arendt-Nielsen, Kevin Hardt, David Manning, A. Vania Apkarian, Paulo Branco

## Abstract

Total knee replacement (TKR) is the gold-standard treatment for end-stage chronic osteoarthritis pain, yet many patients report chronic postoperative pain after TKR. The search for preoperative predictors for chronic postoperative pain following TKR has been studied with inconsistent findings. This study investigates the predictive value of quantitative sensory testing (QST) and PainDETECT for postoperative pain 3, 6, and 12 months post-TKR. We assessed baseline and postoperative (3- and 6-months) QST measures in 77 patients with knee OA (KOA) and 41 healthy controls, along with neuropathic pain scores in patients (PainDETECT). QST parameters included pressure pain pressure threshold (PPT), pain tolerance threshold (PTT), conditioned pain modulation (CPM), and temporal summation (TS) using cuff algometry, alongside mechanical hyperalgesia, and mechanical temporal summation to repeated pinprick stimulation. Compared to healthy controls, KOA patients at baseline demonstrated hyperalgesia to pinprick stimulation at the medial OA-affected knee and cuff pressure on the ipsilateral calf. Lower cuff algometry PTT and mechanical pinprick hyperalgesia were associated with baseline KOA pain intensity. Moreover, baseline pinprick pain hyperalgesia explained 25% of variance in pain intensity 12 months post-TKR and preoperative neuropathic pain scores also captured 30% and 20% of the variance in postoperative pain at 6- and 12-months, respectively. A decrease in mechanical pinprick hyperalgesia from before surgery to 3 months after TKR was associated with lower postoperative pain at the 12 months post-TKR follow-up, and vice-versa. Our findings suggest that preoperative pinprick hyperalgesia and PainDETECT neuropathic-like pain symptoms show predictive value for the development of chronic post-TKR pain.

## Introduction

Knee osteoarthritis (KOA) is the most common progressive multifactorial form of arthritis, affecting over 240 million people worldwide [1] and is characterized by chronic musculoskeletal pain and disability [2]. Total knee replacement (TKR) is the gold-standard treatment for end-stage KOA when non-surgical therapies are no longer effective. Yet, after undergoing TKR, 7 to 34% of patients experience chronic postoperative pain [3, 4]. Given the health and economic burdens of TKR, large efforts have been made to identify risk factors that can predict long-term outcomes, including pain. Meta-analytical evidence supports the role of preoperative pain, other painful body sites, pain catastrophizing, and, to a lesser extent, depression as risk factors for chronic postoperative pain following TKR [5]. However, to date, the predictive value of clinical and psychological features remains insufficient for useful clinical decision-making.

More recently, quantitative sensory testing (QST) has been studied as a possible predictor for TKR success given its ability to directly quantify aspects related to the pathophysiology of chronic pain, including allodynia, hyperalgesia, temporal summation (TS) of nociception, and conditioned pain modulation (CPM) as proxies for sensitization of the nociceptive apparatus [6]. While many KOA patients present QST abnormalities in comparison to healthy individuals [7, 8], including widespread pressure hyperalgesia, facilitated TS, and impaired CPM [6, 9, 10], QST scores can be variable, and this variability has been suggested to be associated with the outcomes of interventions[11]. A recent systematic review supports that preoperative pain pressure threshold (PPT), TS, and CPM are the most frequently associated with chronic postoperative pain. Yet, results are conflicting and difficult to interpret due in part to heterogeneity in QST assessment methods [12].

The presence of neuropathic pain symptoms prior to surgery has also been identified as a potential predictor of postoperative pain after TKR [13]. A widely used tool to assess neuropathic-like pain symptoms in patients with chronic pain is the PainDETECT questionnaire [14]. PainDETECT has been used to subdivide patients with KOA into three groups—non-neuropathic, unclear, and neuropathic pain-like phenotypes—which have been suggested to differentially respond to treatments [15], with the neuropathic pain subgroup also displaying widespread pressure hyperalgesia compared to the other groups [16]. Moreover, higher preoperative PainDETECT scores have also been associated with chronic postoperative pain after TKR surgery [17], and the same study suggests that QST parameters, such as TS and hyperalgesia, can also be explained by neuropathic pain qualities through the PainDETECT questionnaire, raising the question of whether TS and hyperalgesia might be tapping into the same sources of variance.

In summary, QST parameters and neuropathic pain-like qualities are promising potential predictors of TKR outcomes, but more thorough studies are required due to the high variability in methods used [12]. Most studies are correlational, focus only on a single time point, and overlook how preoperative parameters affecting outcomes normalize post-TKR. Lastly, the relationship between QST metrics and neuropathic pain merits further exploration.

The c4, urrent study aimed to i) compare QST parameters between KOA patients and healthy controls, ii) determine if QST parameters and neuropathic pain-like qualities can predict post-TKR pain at 3-, 6- and 12-months follow-up, and iii) assess if changes in QST parameters and neuropathic pain-like qualities post-TKR relate to long-term pain outcomes.

## Methods

### Study Design and Participants

In this observational study, 77 patients (mean age 67.2 years, standard deviation [SD]=6.9, 70.1% female) with chronic KOA awaiting TKR surgery were recruited from Northwestern Memorial Hospital’s Department of Orthopedic Surgery, between July 2019 and November 2022. Patients invited to participate were older than 40 years, met the American College of Rheumatology criteria for KOA, had KOA for more than 6 months, reported knee pain most days of the week for the past month, had a pain intensity score greater than 4/10 at their baseline visit, were scheduled to undergo a TKR surgery within 3 months of consent, were able to read and speak English and were in general good health. Of the 77 patients, 14 had a previous TKR surgery on their other knee (18.2%). A second cohort of 41 age- and sex-matched participants with no history of OA or other chronic pain conditions were recruited as a healthy control group (mean age 65.9 years, [SD]=7.0, 75.6% female). Sample sizes were selected based on an *a priori* power analysis (see below, section *Statistical Analyses*).

Participants were excluded if they i) had evidence of rheumatoid arthritis, ankylosing spondylitis, or other inflammatory arthropathy; ii) reported current recreational drug use or history of alcohol or drug abuse; iii) reported chronic neurologic conditions or psychiatric diseases; iv) were not suitable for MRI scanning; or v) had other significant medical diseases, such as congestive heart failure or peripheral vascular disease. An additional exclusion criterion for healthy controls was ongoing acute or chronic pain. This study is part of a larger project aimed at studying the brain, psychophysical and psychological predictors, and consequences of chronic pain after TKR; in this manuscript, we report the findings for psychophysical QST parameters. The study was approved by the Northwestern University Institutional Review Board (STU00207973), and all participants signed a written informed consent.

Baseline visits with initial assessment procedures were conducted within two weeks of the KOA patient’s TKR surgery. In this first visit, patients consented, completed a series of psychological and clinical questionnaires, underwent a brain scan, and performed a QST assessment. Patients were invited to return for a follow-up assessment at 3 and 6 months following TKR surgery to assess their pain and repeat all procedures. Due to the COVID-19 pandemic, not all patients were able to return to the follow-up visits; the full number of subjects per visit and per analysis is reported whenever appropriate, and the full summary can be found in the supplementary material. Patients were contacted at 12 months for clinical pain assessment only. Healthy control participants completed questionnaires and QST assessments once.

### Pain-related measurements

All patients completed the PainDETECT questionnaire to assess pain intensity and the presence of neuropathic pain-like characteristics. In this questionnaire, KOA patients were asked to report the average pain in the last 4 weeks for their OA knee (VAS, 0–10), which was the main outcome measure at each time point (before TKR, and at 3, 6, and 12 months after TKR). PainDETECT further queries patients regarding their pain pattern, the presence of radiating pain, and the frequency at which they feel characteristic neuropathic pain symptoms, such as burning and prickling [14]. These are quantified to provide a composite score of neuropathic pain profile. If patients did not complete PainDETECT at a given time (n = 6, 3, and 3 for 3, 6 and 12 months after TKR, respectively), we instead collected their pain ratings over the phone.

### Quantitative sensory testing

QST was performed with participants sitting on a reclining chair in a quiet room with their legs extended and slightly bent for comfort. In this study, we used a pinprick stimulator to assess mechanical pain sensitivity and wind-up-ratio (WUR) at the affected knee and a cuff pressure algometry device to measure pressure pain threshold, pain tolerance, temporal summation (TS) and conditioned pain modulation (CPM) at the leg ipsilateral to the knee undergoing TKR. Healthy control participants performed the same QST procedures but with the stimulation laterality chosen randomly.

Pinprick measures of mechanical pain sensitivity and wind-up.

Mechanical pain sensitivity and WUR were assessed on the skin adjacent to the affected knee’s medial border using a pinprick test (see Fig S1). Participants reported their pain rating using a 101-point Numeric Rating Scale (NRS, 0-100) following a single stimulus using a weighted 25.6 g pinprick stimulator. This was first tested on the non-osteoarthritic knee to ensure participants understood instructions and then was performed on the OA knee, while participants were instructed to close their eyes or look away during the stimulation. Subsequently, 10 consecutive monofilament stimulations were applied at one stimulus per second within a 1 cm^2^ area of the skin on the OA knee. After the 10 stimuli, participants reported their average pain intensity. After a brief rest interval, the assessments with a single stimulus and 10 stimuli were repeated once more on the same knee. Pain ratings (mechanical hyperalgesia) for a single prick (prick1) and 10 consecutive pricks (prick10) were registered as the average across the two trials. The ratio between the first and average pain ratings of the 10 stimulations was used to calculate the WUR, which was averaged across the two trials.

#### Cuff Pressure Algometry

To examine dynamic pain responses, TS, and CPM, we used a computer-controlled cuff pressure algometer (Cortex Technology and Aalborg University, Denmark) using a 13-cm wide cuff on the gastrocnemius muscle of each leg. Procedures closely follow previous publications [10, 17]. For the assessment of pressure pain and tolerance thresholds, the cuff was inflated automatically by a computer at a rate of 1 kPa/s until a maximum pressure of 100 kPa was reached. The participants used an electronic visual analog scale (VAS) for continuous recording of pain intensity. Participants were instructed to press a pressure release button when the pain was intolerable. The VAS signal was sampled at 10 Hz, and 0 and 10 cm on the scale were defined as minimum and maximum pain, respectively. The pressure value when the subject rated the pain sensation as 1 cm on the VAS was defined as the pressure pain threshold (PPT). When the subject terminated the exam using the pressure release button, the pressure value was defined as the pain tolerance threshold (PTT).

To assess TS, 10 repeated cuff pressure stimulations with a 1-second duration and 1-second interval between stimuli and a pressure rate of 100 kPa/s were delivered to the affected leg by inflation of the cuff chamber to the PTT recorded during the previous assessment. In the period between stimuli, the cuff was entirely deflated. Pressure at the PTT intensity was used to ensure that subjects perceived the stimulation as painful but not unbearably painful due to the short stimulation time. Subjects rated their pain sensation on the electronic VAS during the cuff inflations, thus not returning to zero between successive stimulations. The VAS score immediately after each stimulus was extracted for TS calculation. The difference between the average of the last 3 VAS scores and the average of the first 3 VAS scores was used to calculate the cuff TS value.

For the CPM assessment, the cuff on the contralateral leg was inflated to a pressure at 70% of the subject’s PTT intensity and held at this constant pressure for the duration of the assessment. The participants were instructed to ignore the stimulus on the contralateral knee and to focus on rating their pain on the electronic VAS for the cuff pressure on their OA knee. Simultaneous reassessment of the subject’s PDT of their OA knee was performed. The difference between PDT during and before the conditioned pain was used to calculate CPM.

### Statistical methods

We specified 7 QST variables of interest from the measures described above, in line with previous studies [12]. From pinprick QST measures, we selected prick1, prick10, and their wind-up ratio (WUR), and from cuff algometry QST measures, we selected PPT, PTT, CPM, and TS. We modeled pain continuously (*i.e.,* through linear modeling) to avoid dichotomizing patients into “chronic pain” or “recovered” based on arbitrary criteria [18]. Linear model residuals were right-skewed, partly because many patients reported relatively minor pain (NRS < 3/10, see below) at 6 and 12 months after surgery. Therefore, we log-transformed all pain scores (*i.e.,* pain_log_ = log_10_(pain+1)), which yields normally distributed residuals. In addition, we log-transformed the WUR since it is a ratio, converting it from a multiplicative construct to an additive one. Hence, prick1 and prick10 were first log-transformed, and then their ratio was calculated to obtain WUR. We computed *post hoc* correlations using the nonparametric Spearman’s. For cross-sectional comparisons between KOA patients and healthy controls, an analysis of covariance (ANCOVA) was performed by comparing how QST outcome differed between groups while controlling for age and sex. We adjusted the *p*-values to control for the false discovery rate (FDR).

For the longitudinal predictive analyses, we probed the variables deemed abnormal in the cross-sectional analyses (i.e., compared to healthy controls) and assessed their predictive ability; other variables were tested *post hoc* as exploratory analyses. We leveraged the repeated pain assessments by modeling the outcomes across time using generalized estimating equations (GEE)—this estimates “population effects” (cf. mixed-effects models) while effectively controlling for the covariance between repeated measures. Pain ratings at baseline, 3, 6, and 12 months after surgery were predicted from baseline QST parameters, again controlling for age and sex. We adjusted for multiple comparisons across time points using the multivariate *t* approach (*i.e.,* ‘mvt’ in the R package *emmeans*), which adjusts *p*-values based on the multivariate *t* distribution of the model, hence taking into account the covariance structure of the data. Finally, measures showing statistically significant (adjusted *p* < 0.05) predictive ability were further assessed longitudinally—how they change or normalize after surgery. To do so, we fit a multivariable linear model predicting pain 12 months after TKR with baseline and change in QST scores (i.e., change is 3 months minus baseline, or 6 months minus baseline), again with age and sex as covariates of no interest. For these analyses, two models were run separately for 3 and 6 months due to missing data, as the patients at 3 and 6 months do not fully overlap.

Sample sizes were chosen based on an *a priori* power analysis; however, the actual numbers collected deviate slightly from initial estimates due to the COVID-19 pandemic restrictions. Power was calculated using G*Power 3.185 and the R package pwd. For the cross-sectional study, it was estimated that a minimum of 30 participants per group was sufficient to detect an effect of *d* = 0.8 at 80% power, which was deemed reasonable given our preliminary data. For the longitudinal prediction analyses, we initially calculated the power to classify chronic versus recovered patients using logistic regression and estimated to need to collect 105 patients, translating to a statistical power of 82% for variables with odds ratios > 2. As pain ratings did not conform to a bimodal distribution (see above) and the numerous previously described issues with dichotomizing outcomes [19], we instead ran linear regressions which also confers more statistical power [19]. The power for this analysis was thus not explicitly calculated *a priori*; *post hoc* power can be extrapolated from the *p*-values [20]. Finally, we estimated the need to collect 30 subjects for the within-subject comparison at 80% power for the longitudinal change analyses, assuming medium-sized effects (*r* among repeated measures = 0.6 or 36% shared variance). We initially also planned to have a validation dataset to minimize type I errors, but that proved unfeasible due to the difficulty of recruiting patients during COVID-19 restrictions. Statistical power notwithstanding, here we report *all* results and transparently report measures of uncertainty (CIs), based on which readers can make conclusions about which effects the study could estimate well—if large effects *and* zero. Effect sizes are also reported whenever appropriate, using R-squared for linear regressions and are contained within the CI, then no strong conclusions can be made due to the poor precision of the estimate Cohen’s d for group comparisons, i.e., the average of one group minus the other, divided by the pooled standard deviation. Whenever analyses are performed on log-transformed data, we calculated R^2^ 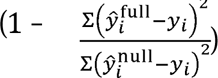 and adjusted R^2^ 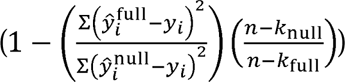 based on *y_i_* and 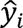 transformed back to their raw scales, where *n* is the number of participants and *k* is the number of parameters, including the intercept. These R^2^ values thus account for any bias associated with fitting models on the log scale.

## Results

### Preoperative QST profiles in KOA and pain-free individuals

For measures collected with the cuff algometer, participants with KOA showed significantly lower PTT compared with healthy controls (*t*(104) = −2.40, *p* = 0.018, *p*_FDR_ = 0.047, *d* = 0.48, Fig. 1A). All other cuff assessment measures did not statistically significantly differ between groups (all *ps* > 0.15, Fig. 1A). For measures collected with the mechanical pinprick stimulator, KOA patients reported higher pain than healthy controls for both the single prick (prick1, *t*(97) = 4.02, *p* < 0.001, *p*_FDR_ < 0.001, *d* = 0.83) and the consecutive 10 pricks (prick10, *t*(97) = 4.19, *p* < 0.001, *p*_FDR_ *<* 0.001*, d* = 0.87). WUR did not show statistically significant differences between groups (*p* = 0.73). Together, our findings suggest that KOA patients show mechanical hyperalgesia in the knee – and a smaller effect of pressure pain tolerance at the calf – yet otherwise normal temporal summation and conditioned pain modulation compared to pain-free individuals.

**Figure 1.**
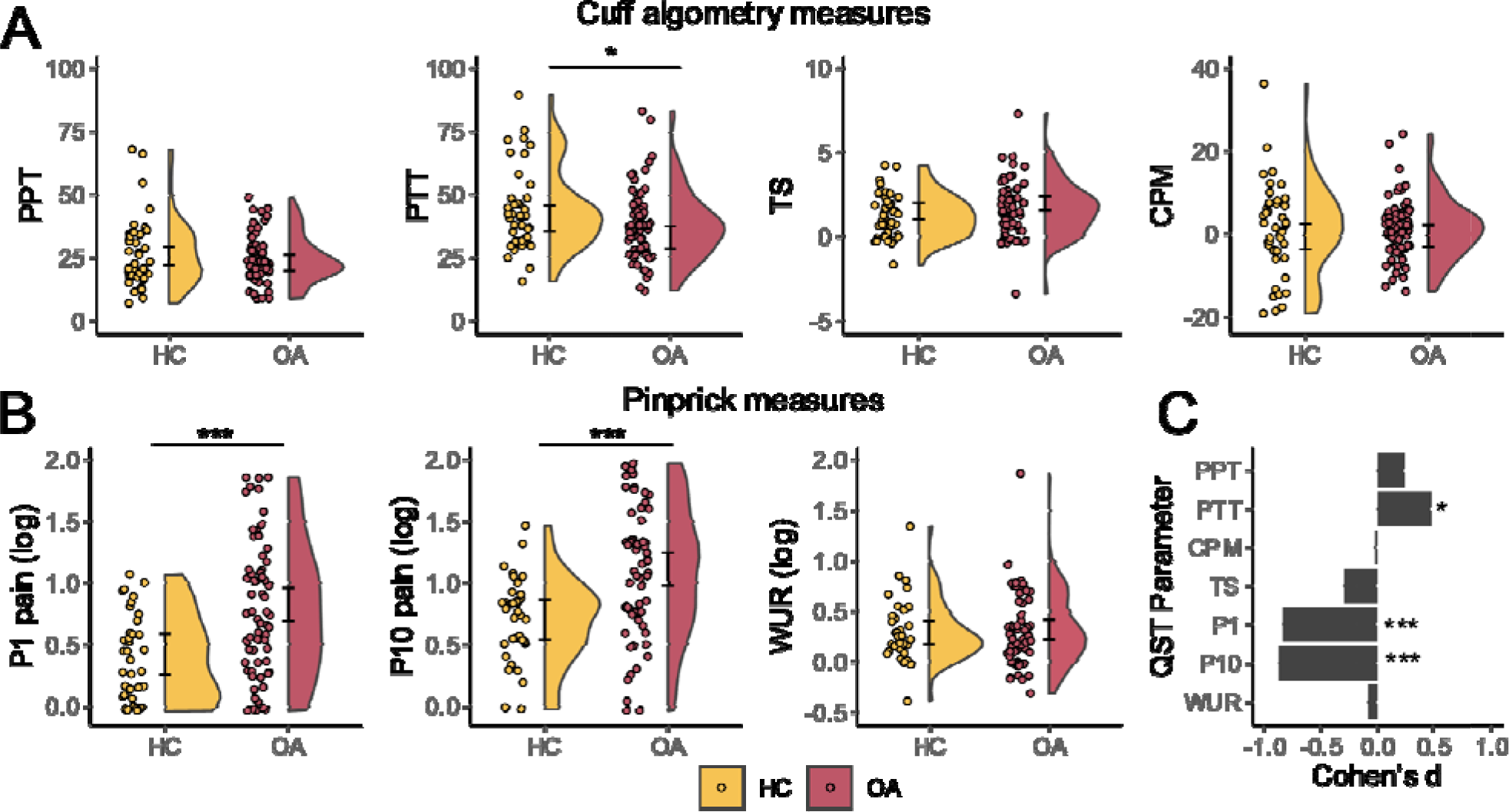
Knee osteoarthritis (KOA) patients show pronounced mechanical hyperalgesia before surgery but otherwise similar quantitative sensory testing parameters (QST). A) for cuff algometry measures, only PTT was significantly different between healthy controls (HC) and knee osteoarthritis (OA) groups, with smaller values for KOA. B) KOA patients rated both the single pinprick (P1) and the consecutive 10 pinpricks (P10) as more painful than healthy controls (HC). C) Cohen’s *d* effect size for each QST parameter shows large effect sizes for pinprick measures, and small for pain tolerance threshold (PTT). * *p* < .05; *** *p* < .001, after FDR correction. 95% confidence intervals are reported at the edge of the violin plots. CPM, Conditioned pain modulation. TS, temporal summation. WUR, wind-up-ratio. PPT, pressure pain threshold, PTT: pressure tolerance threshold.

### Predicting pain after total knee replacement with QST

Out of the 77 patients enrolled in the study, 54 patients provided pain ratings at the 3-month mark, 35 patients at the 6-month mark, and 46 patients at the 12-month mark – the endpoint of the study.

Patients’ pain improved substantially after surgery; however appreciable variability in pain ratings across subjects is observed: at baseline, patients reported an average pain of 6.3/10 ([SD]=2.1), 2.5/10 at 3-months ([SD]=2.1), 1.7/10 at 6-months ([SD]=1.9) and 1.9/10 at 12-months ([SD]=2.4), see Figure 2A. We first assessed if pain ratings registered during pinprick stimulation and PPT–– the three abnormal measures found in the cross-section analysis – were associated with pain before or at 3, 6, and 12 months after TKR. The pain reported with the single prick was associated with baseline pain (β = 0.105, *p* < 0.001, *p_mvt_* < 0.001), and it predicted pain at 12 months (β = 0.243, *p* = 0.002, *p_mvt_* = 0.008). Single pinprick measures were initially found to be associated with pain at 3 months (β = 0.131, *p* = 0.042) and 6 months after TKR (β = 0.166, p = 0.055), with confidence intervals largely supporting positive associations with pain (Figure 2B), but these associations did not remain statistically significant after adjusting for multiple comparisons (3 months: *p*_mvt_ = 0.14; 6 months: *p*_mvt_ = 0.18). Model fits depict the positive association between baseline prick and clinical pain preoperative and at later time points (Figure 2C). Baseline prick was able to explain 13% of variance in pain ratings at baseline, 9% at 3 months, 12% at 6 months, and 25% at 12 months, thus suggesting that mechanical hyperalgesia may account for a significant portion of the variance in chronic postoperative pain one year after TKR, long after the tissue has healed.

**Figure 2.**
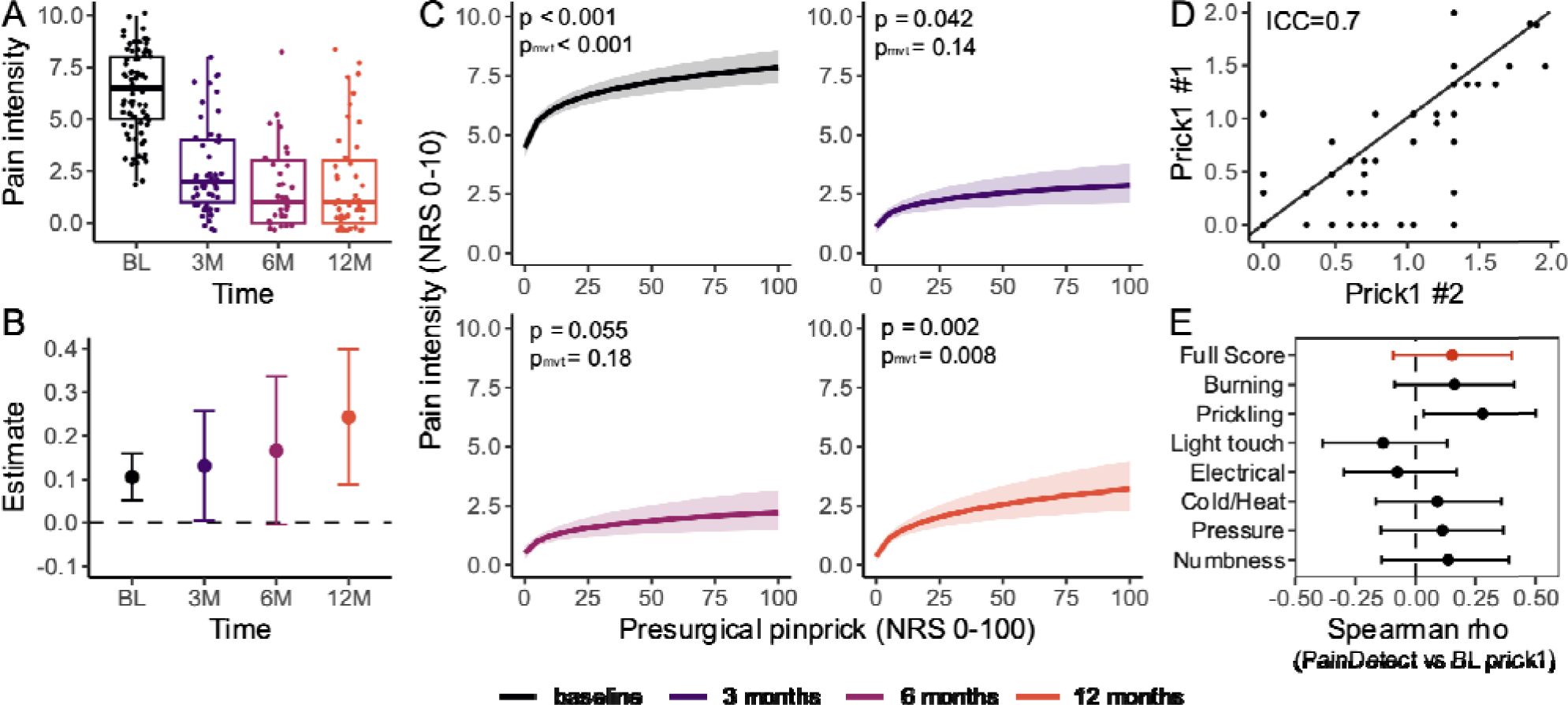
Pinprick stimulation predicts long-term pain outcomes. (A) Pain after surgery improves substantially, yet with large variability in outcomes. (B) Higher pinprick pain at baseline is predictive of higher pain at all time points. Solid line reflects the model fits, with corresponding 95% confidence intervals (CIs), with the largest and most significant effect being the prediction of pain at 12 months after TKR. (C) Model predictions and CIs for postoperative pain at each time point for respective single-pinprick values. D) Pearson correlation shows that the two pinprick trials yield similar results, thus showing robustness of the experimental procedure. (E) Spearman correlations between baseline pinprick measures and pain characteristics as measured with *PainDETECT* show that subjects with higher pinprick pain more frequently report sensations of tingling and prickling on the affected knee. ICC, Intraclass coefficient. NRS, Numeric Rating Scale.

Pain ratings for ten consecutive pinpricks were also significantly associated with baseline pain (β = 0.085, *p* = 0.004, *p*_mvt_ = 0.013). While this measure also predicted pain at 12 months (β = 0.196, *p* = 0.026), this result was not statistically significant after adjusting for multiple comparisons (*p*_mvt_ = 0.081, confidence intervals and model can be inspected in Supplementary Fig. 1). Pain ratings for the ten consecutive pinprick values were not statistically significantly predictive of pain at either 3 months (β = 0.049, *p* = 0.479, *p*_mvt_ = 0.872) or 6-months after TKR (β = 0.142, *p* = 0.075, *p*_mvt_ = 0.218).

Finally, baseline PTT values were not statistically significantly associated with pain at either timepoint: βs = 0 (95% CI: −0.003, 0.003), 0.001 (95% CI: −0.004, 0.007), 0.002 (95% CI: −0.005, 0.008), and 0.001 (95% CI: −0.003, 0.006) for baseline, 3 months, 6 months and one year, respectively, all *ps* > .50).

For completion, we also explored, post hoc, the predictive ability of the remaining QST measures. PDT, CPM, and WUR were not statistically significantly associated with baseline pain or able to predict pain at future time points (all *ps* > .42, see Supplementary Material). TS assessed with cuff algometry was associated with baseline pain (β = 0.025, *p* = 0.021 *p*_mvt_ = 0.072) yet this result did not survive adjustment for multiple comparisons. Further, TS was unable to predict pain at either of the three postoperative time points (all *ps* > 0.196).

### Measurement Properties of Pinprick

We further explored these findings with additional analyses. First, we assessed how prick 1 and prick 10 independently account for variance in 12-month pain outcomes. A Pearson correlation showed that these measures were strongly correlated with each other (*r* = 0.85, *p* < 0.001). When the two were added to a multivariable model, the model favored prick 1 values (β = 0.35 ± 0.17) over prick 10 values (β *=* −0.11 ± 0.19). Since prick 1’s variance seems to be favored and prick 10 predictions did not survive adjustment for multiple comparisons, here, we only report the results from prick 1 measures for further analyses. Second, given that pinprick was performed twice (i.e., two trials), we assessed the test reliability of the two pinprick ratings. This entailed calculating the intraclass correlation (ICC, two-way mixed, absolute agreement) between the first and second trials, which showed good test-retest reliability with an ICC = 0.70 (Fig 2D). Second, since mechanical hyperalgesia and tactile allodynia are hallmark symptoms of neuropathic pain [21], we explored associations between baseline pinprick scores and the frequency and characteristics of each painful sensation as measured by painDETECT questionnaire, to assess if patients with higher evoked pain with pinprick also report other abnormal sensations in the knee. For the single pinprick measures, Spearman rank-based correlations showed that pinprick pain ratings significantly correlated with the frequency upon which patients report “prickling sensations” in the osteoarthritic knee (ρ = 0.28, 95% CI: (0.04, 0.50), *p* = 0.028, Figure 2E). No additional significant associations were found for any other sensation measured with PainDETECT, including the total score (ρ = 0.15, *p* = 0.24). For prick10, we again found associations between pinprick scores and prickling pain (ρ= 0.27, *p* = 0.03) and the full PainDETECT score (ρ = 0.30, *p* = 0.016). No additional statistically significant associations were found.

### Predicting pain after total knee replacement with neuropathic pain characteristics

Given previous findings showing that neuropathic pain characteristics predict TKR outcomes (Kurien et al., 2018), we further examined if baseline neuropathic scores provided by PainDETECT could also predict future pain outcomes. The distribution of PainDETECT scores shows patients have significant variability in neuropathic pain qualities (Fig 3A). PainDETECT scores were associated with baseline pain (β = 0.008, *p* < 0.001, *p_mvt_* < 0.001), not statistically associated with pain at 3 months (β = 0.012, *p* = 0.052, *p*_mvt_ = 0.162) but significantly predictive of pain at 6 and 12 months after TKR (β = 0.024, *p* < 0.001, *p*_mvt_ = 0.002, and β = 0.021, *p* = 0.004, *p*_mvt_ = 0.014, respectively, see Figure 3B for confidence intervals and Figure 3C for model predictions). For all time points, higher preoperative PainDETECT scores, which measure neuropathic pain characteristics, were associated with greater pain after TKR. This model was able to explain 16% of variance in pain ratings at baseline, 12% at 3 months, 30% at 6 months, and 20% at 12 months. To further explore this finding, we assessed the correlation between each pain characteristic from PainDETECT and pain at 12 months, and we found that higher chronic postoperative pain was correlated with increased frequency of burning sensations (ρ = 0.46, 95% CI: (0.17, 0.69), *p* = 0.002), tingling or prickling sensations (ρ = 0.43, 95% CI: (0.14, 0.68), *p* = 0.004), pain with cold or heat stimuli (ρ = 0.31, 95% CI: (0.002, 0.60), *p* = 0.043) and numbness in the area (ρ = 0.41, 95% CI: (0.12, 0.65), *p* = 0.006), see Figure 3D.

**Figure 3.**
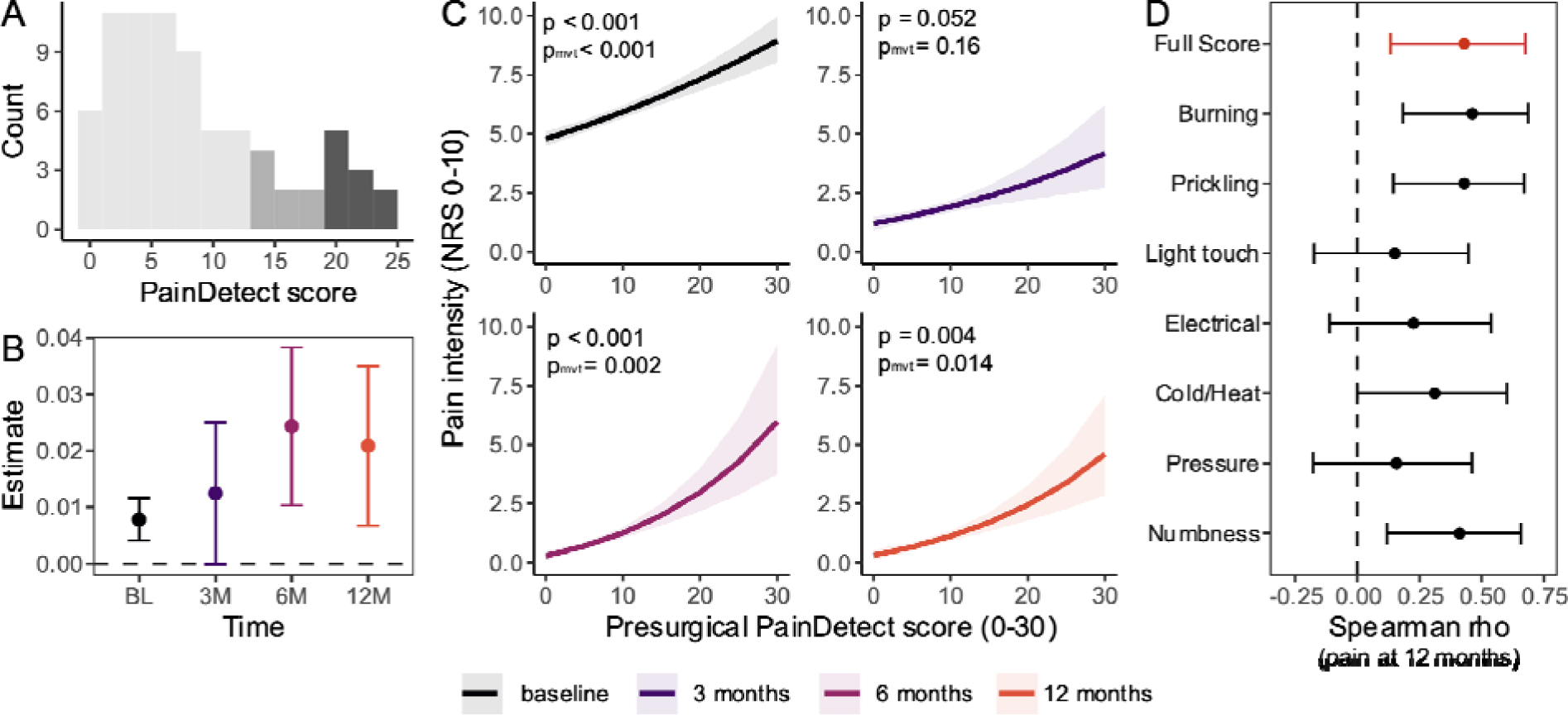
Higher PainDETECT scores are also associated with worse long-term pain outcomes. A) PainDETECT scores show large variability, with 40 subjects falling in the “unlikely to have a neuropathic component” category, 12 with “uncertain” category, and 10 likely having a neuropathic pain component. B) Estimates and confidence intervals for the prediction at four time points show that baseline *PainDETECT* is significantly predictive of pain at baseline, 6 months, and 12 months. The results are stronger and less compatible with the null for 6 and 12 months after TKR. (C) Model fits and corresponding 95% confidence intervals for each timepoint, again supporting that higher neuropathic pain characteristics are associated with higher postoperative pain. D) Spearman correlations between pain at 12 months, and each of the pain sensations assessed with PainDETECT show that prickling and tingling, numbness, and burning sensations in the osteoarthritic knee at baseline are correlated with the pain patients report 12 months after TKR surgery.

### Modelling hyperalgesia and neuropathic symptoms

Given the previously reported ability of pinprick hyperalgesia and PainDETECT scores to predict postoperative pain, we assessed whether these capture the same or different sources of variation in pain outcomes. Further, we also tested if the predictive ability of these measures is better explained by baseline pain. To do so, the two most significant parameters from the above analyses (prick1 and PainDETECT score) were singled out and, together with baseline pain, age, and sex, added to a single multivariable linear model predicting pain intensity 12-months after TKR. Both prick1 (β = 0.21, 95% CI: (0.02, 0.41), *p* = 0.030) and PainDETECT score (β = 0.02, 95% CI: (0, 0.03), *p* = 0.046) were independently associated with pain at 12 months, while baseline pain (β = 0.14, 95% CI: (−0.57, 0.80), *p* = 0.69), age (β = 0, 95% CI: (−0.01, 0.02), *p* = 0.71) and sex (β = −0.07, 95% CI: (−0.33, 0.19), *p* = 0.56) were not statistically significant. Relative importance metrics [22] showed that prick1 accounted for 15% unique variance in pain outcomes (log scale), PainDETECT for 13% of variance, baseline pain 4%, and age and sex 0%, thus supporting that prick1 and PainDETECT scores do capture unique sources of variance in clinical pain one year after TKR.

### Changes in hyperalgesia and long-term outcomes

Finally, patients returned for QST assessment 3 and 6 months after surgery. Thirty-six patients returned at 3 months and 25 at 6 months. A linear model with baseline prick values and (i) the change from baseline to 3 months or (ii) the change from baseline to 6 months was used to predict pain at 12 months. This model aimed to assess if changes in the abnormal QST parameters (i.e., their persistence or reversal) can further relate to long-term outcomes. At three months, this two-parameter model shows that both baseline values (β = 0.545, 95% CI: (0.29, 0.80), *p* < 0.001) and their change (β = 0.38, 95% CI: (0.11, 0.65), *p* = 0.008) predict the outcome. More specifically, greater preoperative prick pain (Fig. 4a) was associated with greater long-term pain, even after adjusting for change in these parameters. Further, greater decreases in pain reported during pinprick stimulation predicted lower long-term clinical pain after adjusting for baseline pain (Fig. 4b). In other words, improvements in mechanical hyperalgesia (i.e., negative change) were associated with less clinical pain after 12 months, and vice-versa (Fig. 4c). Together, the full model explained 67% of the variance in 12-month pain (*F*_4,19_ = 5.50, *p* = 0.004, R^2^ = 0.67, adj. R^2^ = 0.63). Surprisingly, this result was not observed for the change at 6 months post-surgery: while baseline parameters were still predictive of outcomes (β = 0.39, 95% CI: (0.03, 0.75), *p* = 0.036), their change from baseline to 6 months was not statistically significant (β = 0.03, 95% CI: (−0.40, 0.46), *p* = 0.89). The full model at 6 months did not significantly predict pain at 12 months (*F*_4,13_ = 2.02, *p* = 0.15, see Fig 4d-f).

**Figure 4.**
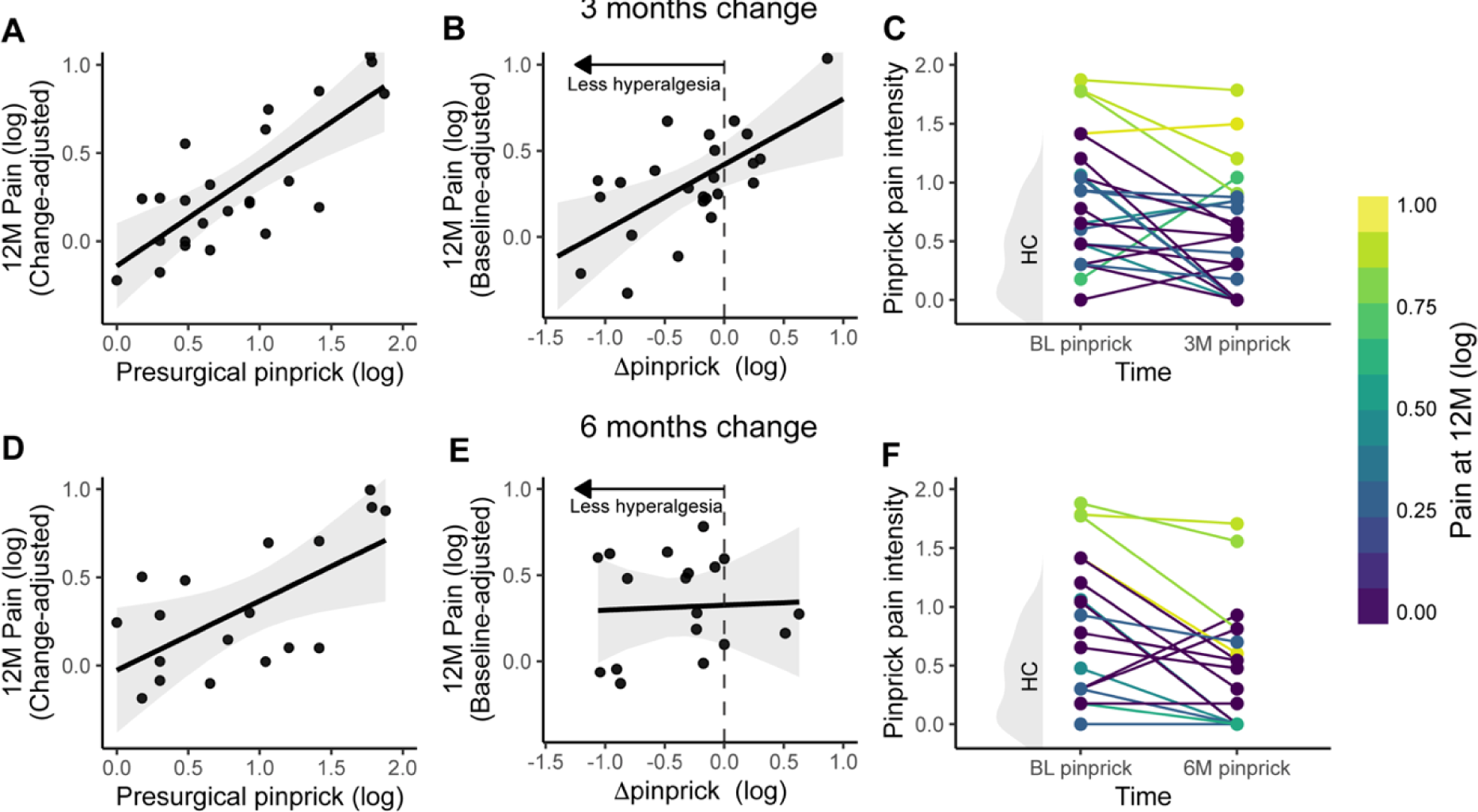
Normalization of mechanical hyperalgesia 3 months after TKR is related with long-term outcomes. Mechanical hyperalgesia changes following TKR. For patients whose data was collected preoperatively and at three months, baseline pinprick pain is associated with worse outcomes when controlling for the change (A), and change from preoperative to three-months pinprick pain is also associated with long-term outcomes when controlling for baseline pinprick measures (B). Line plots of repeated measurements illustrate this association, with patients with normative pinprick pain values, or those who return to normative values after surgical intervention. Healthy control (HC) distribution for pinprick pain intensity is highlighted in gray on the left axis) have low pain 12 months after surgery. In contrast, patients with high preoperative hyperalgesia, or whose hyperalgesia worsened to abnormal states, report the highest pain after 12 months (C). The same analyses for 6 months show again that baseline pinprick pain ratings is associated with pain at 1 year when controlling for the change(D), yet the change in parameters from preoperative to six-month pinprick pain is not significantly related to 12-month pain (E). Individual subject trajectories for baseline and 6 months pinprick values can be seen in panel F.

## Discussion

The current study found that KOA patients showed mechanical hyperalgesia and decreased PTTs, but otherwise, normal QST parameters compared to age- and sex-matched healthy controls. Preoperative pinprick hyperalgesia and neuropathic pain-like qualities (as assessed by PainDETECT) were predictive of presurgical and one-year postoperative pain intensities. Preoperative pinprick pain intensity and neuropathic pain-like attributes independently predicted postoperative pain. Further, changes in pinprick pain ratings from baseline to three months also predicted one-year pain outcomes, with improvements in hyperalgesia at 3 months predicting less pain one year later, and together with baseline pain, this model captured 67% of the variance in postoperative pain at 12 months. The current findings indicate that pinprick hyperalgesia on the OA-affected knee is associated with greater chronic postsurgical pain.

### OA patients show hyperalgesia but otherwise normal QST parameters

A growing body of evidence suggests that patients with OA exhibit various site-specific or widespread sensory abnormalities [6, 7], which are thought to reflect increased gain in nociceptive pathways. In this study, patients with KOA reported higher levels of evoked pain in response to both a single pinprick and a series of ten consecutive pinpricks and had lower cuff PTT as compared to healthy controls. KOA is often associated with pressure hyperalgesia when compared to healthy pain-free individuals (for a review, see [23]). Studies using pinprick assessments also find that KOA patients present hyperalgesia at the skin over the affected knee [24, 25], and that the magnitude of this hyperalgesia is associated with OA disease severity [24]. Hyperalgesia to mechanical stimuli is a common indicator for peripheral sensitization, and potentially also for sensitization at the spinal cord level, given the chronic and ongoing barrage of nociceptive signaling in OA [26–28]. We observed significant hyperalgesia at the medial patella and, to a lesser degree, the ipsilateral calf. These areas overlap in their respective dermatomes and correspond to the same spinal segments [29], which could indicate the involvement of central sensitization. On the other hand, the differences in effect sizes for prick and pressure stimuli are notable, with more pronounced hyperalgesia near the lesion site. Besides location, pressure stimuli primarily target C-fiber-rich deep tissue nociceptors, whereas prick stimuli also target skin-level A-delta nociceptors [30–32], so further studies are needed to understand these differences and the relative contribution of central vs. peripheral mechanisms.

No statistically significant differences were found between KOA patients and healthy controls for all other collected QST parameters, including TS, which is traditionally regarded as a marker of central sensitization, and CPM, here taken as a proxy for descending modulation. Previous studies have shown that KOA patients exhibit facilitated TS and impaired CPM [6, 9, 17]. However, findings in this field remain equivocal [33, 34]. Our results could suggest that effect sizes for these measures are small; thus, studies with small-to-medium sample sizes may have low power, yielding results that fall on either side of the threshold for “statistical significance.” Alternatively, it could also suggest that CPM and TS abnormalities are not universally present in OA patients and are only found in a subset of patients, possibly as a consequence of disease severity, duration, and other underlying factors, in line with recent proposals for the existence of multiple osteoarthritis phenotypes stemming from different pathological sources [8, 35]. However, in our sample, KOA’s distributions were unimodal and relatively homogeneous in comparison to healthy controls (Figure 1).

### Mechanical pain intensity scores predict long-term pain outcomes

Our study showed that higher mechanical hyperalgesia at baseline, specifically prick1, is associated with higher baseline pain and is predictive of postoperative outcomes at later time points. This finding has important translational significance, as the prediction is strongest for pain at 12 months after surgery when the healing process should be complete. The high reliability (ICC) observed among pinprick trials in our study, combined with the predictive ability of each trial when assessed independently, further reinforces the robustness of our findings. There is some evidence that preoperative PPTs measured at the site of injury can predict long-term pain outcomes (see [36] for a review), supporting the idea that abnormal preoperative nociceptive processing is associated with the persistence of pain after surgery. Comparatively fewer studies have looked at preoperative von Frey or pinprick pain intensity ratings to predict postoperative pain after total joint replacement: one study found that pain intensity reported during von Frey assessment prior to TKR can predict pain 2 days after surgery [37], but not pain with range of motion 6 months after surgery [38], and for total hip replacement, no associations were found between presurgical pinprick intensity and pain 6 weeks after total hip replacement [39]. Future studies are needed to clarify the generalizability of our findings.

We did not find any other preoperative QST parameters to be statistically significantly associated with pre-TKR pain levels nor predictive of postoperative pain. Other studies have found high TS and impaired CPM to be predictive of postoperative pain intensity after TKR surgery [17, 40–48]. Yet, it is important to note that there are again largely inconsistent results regarding the predictive value of QST, exacerbated by heterogeneous methodologies [12, 36]. Furthermore, in our sample, none of the above measures appreciably deviated from the healthy controls in the first place, suggesting they were not “abnormal”. This could also play a role in our null results – perhaps these measures are only predictive in subgroups of patients showing abnormal CPM or TS.

### Neuropathic-like symptoms also predict long-term outcomes

It is well-established that sensory abnormalities, including allodynia and hyperalgesia, are hallmark symptoms of neuropathic injury, as widely demonstrated in the spared nerve injury rodent model [49] and in several neuropathic pain conditions such as diabetic polyneuropathy [28, 50]. Although historically, OA has been regarded as a model of nociceptive pain resulting from joint tissue damage and inflammation, extensive data challenges this notion [2, 29, 51], and indeed OA patients frequently report neuropathic pain symptoms [16, 52, 53]. Further, OA patients with higher neuropathic pain symptoms also present mechanical hyperalgesia [16], suggesting a link between the two. Perhaps most importantly, the presence of neuropathic pain characteristics has also been put forward as a predictor of chronic postsurgical pain after TKR [13, 54], and a previous study suggests that painDETECT scores can predict long-term pain outcomes after TKR above the predictive value of hyperalgesia [17]. Our data suggest that preoperative neuropathic pain-like symptoms, as assessed by the PainDETECT score, were predictive of postoperative outcomes at 6 and 12 months after surgery. Further, when we examined painDETECT individual items individually, the most predictive sensation was prickling. Importantly, prick10 pain ratings (and to a lesser extent prick1 pain ratings) were correlated to the painDETECT scale, which could be construed as pinprick and painDETECT values reflecting similar sources of variance (i.e., tapping into the same underlying mechanisms). To test this explicitly, while also controlling for preoperative pain, we built a multivariable model with these three parameters and found that neuropathic pain qualities and hyperalgesia explain unique variance in pain outcomes, while baseline pain is of lesser importance in the presence of these two parameters. This suggests that measured hyperalgesia and neuropathic pain qualities should be considered separate factors when attempting to predict treatment outcomes, potentially reflecting synergistic but independent mechanisms.

### Normalization of hyperalgesia after surgery is associated with long-term outcomes

We followed patients after TKR to understand how abnormal QST parameters at 3 and 6 months after surgery would relate to long-term outcomes. Previous studies have shown that abnormal PPTs to hand-held pressure algometer and impairment of CPM tend to normalize after TKR if the surgery provides pain relief [55]; this normalization might be associated with better outcomes after surgery [40]. Our findings support these notions, with the change in pinprick intensity from baseline to 3 months after TKR correlating with the pain reports at 1 year after TKR, even when controlling for presurgical pain. Importantly, presurgical pinprick pain remained a statistically significant predictor of long-term outcomes when controlling for its change too, suggesting that the mechanisms underlying abnormal hyperalgesia at baseline also play an important role in chronic postsurgical pain regardless of the normalization, possibly due to the fact patients with high hyperalgesia are also the ones less likely to change. Surprisingly, this result was not consistent when assessing changes at 6 months. With sample size limitations in mind, we have two speculations. First, it could be the case that at 3 months following surgery, some patients may still have abnormal sensitivity of the knee as they are still recovering from the tissue insult caused by the surgery, which could have influenced these results. Second, this result could also suggest an ongoing process where postoperative pain over time is less dependent on peripheral mechanisms and more reliant on distinct neuroplastic properties, particularly in the brain [56]. Future analyses examining brain parameters and their changes at these time points may differentiate between these options.

### Limitations

This study presents some limitations. First, data collection was conducted during the COVID-19 pandemic, which impacted retention rates and might also have played a role in patients’ pain outcomes. Furthermore, we did not instruct our patients with KOA to discontinue their use of analgesic pain medication before their first research appointment, potentially impacting their pain perception thresholds and sensitivity to pain. Third, the small sample size precludes us from developing unbiased predictive models (i.e., out-of-sample prediction) with rigorous validation. Future large-scale studies using measures of neuropathic profiles and mechanical pain sensitivity are needed to ascertain their real-world predictive value for TKR success (model validation), and further, studies to directly investigate the clinical value of implementing prediction models (model utility). Finally, our sample was 70% female, in line with the known higher prevalence of knee OA in females; we adjusted for sex in all analyses, but due to lack of statistical power to study sex main effects and interactions[57], we did not study sex-specific effects.

## Conclusion

In summary, the present study shows that mechanical pinprick and cuff-algometry hyperalgesia and neuropathic pain-like qualities are relevant for estimating postoperative pain following TKR surgery. Moreover, how these parameters change following surgery, especially in relation to patients’ clinical pain, may provide important mechanistic insight into variable outcomes following TKR. Further work is needed to rigorously validate clinical prediction models and assess their clinical utility.

## Data Availability

All data produced in the present study are available upon reasonable request to the authors

## Acknowledgments

The authors would like to thank all members of the Apkarian lab for their feedback on the manuscript. This work was supported by the National Institutes of Health grants to AVA: P50 DA044121 and grant R01AR074274. Center for Neuroplasticity and Pain (CNAP) is supported by the Danish National Research Foundation (DNRF121). The Center for Mathematical Modeling of Knee Osteoarthritis (MathKOA) is funded by the Novo Nordisk Foundation (NNF21OC0065373).

