## Supplementary Material for "Mechanical hyperalgesia and neuropathic pain qualities impart risk for chronic postoperative pain after total knee replacement"

Table 1. Data collected at baseline, for osteoarthritis patients (OA) and healthy controls (HC)

| Measure | OA | HC |
| --- | --- | --- |
| PPT | 65 | 41 |
| PTT | 67 | 41 |
| CPM | 64 | 41 |
| TS | 62 | 41 |
| Prick1 | 64 | 37 |
| Prick10 | 64 | 37 |
| WUR | 64 | 37 |

Table 2. Number of subjects with data collected at baseline and pain ratings at follow-up.

| Measure | Baseline pain (total ratings: 76) | 3 months  (total ratings: 54) | 6 months  (total ratings: 35) | 12 months  (total ratings: 46) |
| --- | --- | --- | --- | --- |
| PPT | 64 | 48 | 33 | 40 |
| PTT | 66 | 50 | 35 | 42 |
| CPM | 64 | 48 | 33 | 39 |
| TS | 62 | 45 | 31 | 37 |
| Prick1 | 64 | 46 | 31 | 38 |
| Prick10 | 64 | 46 | 31 | 38 |
| WUR | 64 | 46 | 31 | 38 |
| PainDETECT | 76 | 53 | 34 | 45 |

Table 3. Number of subjects with longitudinal QST (prick 1) assessments and long-term outcomes (prick 1 + 1 year outcome)

| Measure | Baseline and 3 months | Baseline and 6 months |
| --- | --- | --- |
| Prick 1 | 36 | 25 |
| Prick 1 + 1 year outcome | 24 | 18 |

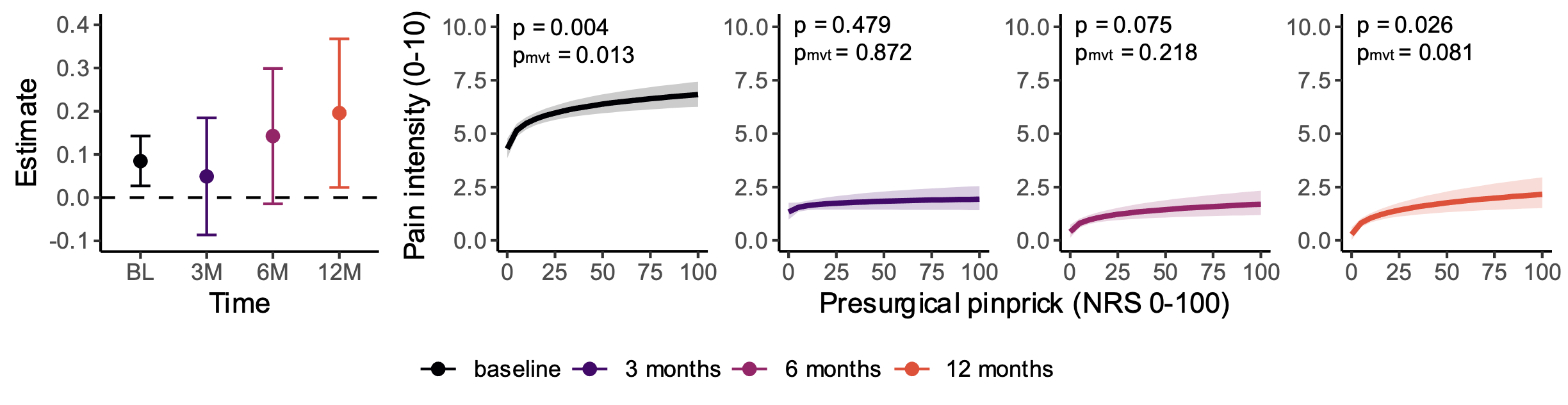

**Figure S1. Prick10 pain intensity predicts baseline pain, and marginally pain 12 month after surgery.** *Higher pinprick pain at baseline is predictive of higher pain at baseline, and marginally at 12 months. Solid line reflects the model fits, with corresponding 95% confidence intervals (CIs), with the largest and most significant effect being the prediction of pain at baseline. (C) Model predictions and CIs for postoperative pain at each time point for respective prick10 values.*

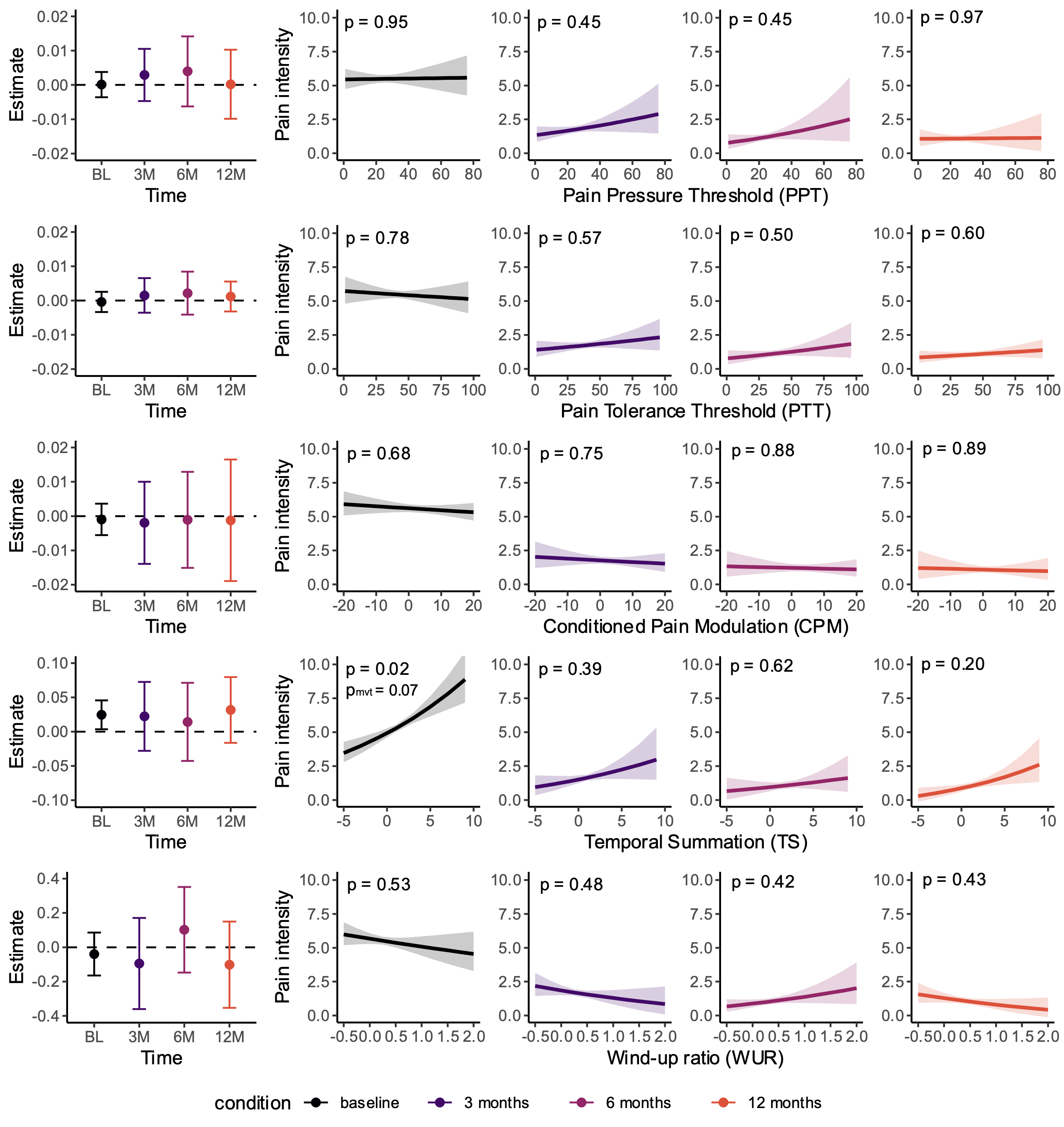

**Figure S2. Predictions for all other QST parameters.** Left panel represents the model fits and corresponding 95% confidence intervals for each timepoint, right panel shows the predictions at each timepoint. All analyses adjusted for age and sex. The only significant effect was temporal summation predicting baseline pain (p = 0.02), which did not survive multiple comparisons (p*_mvt_* = 0.07).
